# Implementing a Resilience and Compassion Program: Students’ Perceptions in Rural Colombia

**DOI:** 10.1101/2025.08.19.25333695

**Authors:** Lina Maria Gonzalez Ballesteros, Camila Andrea Castellanos-Roncancio, Oscar Eduardo Gómez Cárdenas, Isabela Osorio Jaramillo, Luis Eduardo Mojica, Jennifer Clavijo-Marín, Luis Alberto López-Romero

**Author notes:** Corresponding Author: Lina María Gonzalez Ballesteros, MD, PhD(c) Fundación Saldarriaga Concha, Bogotá, Colombia.

## Abstract

In low- and middle-income countries such as Colombia, school-based mental health interventions that promote resilience and compassion have been consolidated as effective strategies to promote emotional well-being in adolescents, especially in contexts affected by the armed conflict. “Conmigo, Contigo y Con Todo” (3C) is a psychoeducational model designed to strengthen resilience, compassion and inclusion in school settings, through strategies based on cognitive-behavioral and third-generation approaches. This qualitative study aimed to explore children and adolescents’ perceptions of resilience, compassion and inclusion, and to understand how these were transformed after their participation in the 3C model and three months later, compared to their initial perceptions. A total of 335 focus groups were conducted with students aged 6 to 18 years before, during and after the intervention. Data were analyzed using a six-phase thematic analysis with inductive coding. Findings evidenced a dynamic process in which participants re-signified resilience as a proactive ability to cope with adversity, while compassion and empathy evolved into concrete actions of mutual support and care. This process reflected the practical integration of the model in their school and personal contexts. The results underscore the potential of culturally sensitive school-based interventions to promote psychosocial well-being, highlighting the importance of strategies that empower students as active agents in their emotional and social development.

## Introduction

Mental health disorders, such as anxiety and depression, are increasingly prevalent among children and adolescents in Latin America, and this situation coexists with a wide gap—estimated between 64% and 86%— in access to treatment that reflects the inadequate attention to their mental health needs (Kohn et al., 2018). In Colombia, the prevalence of at least one mental disorder in children aged 7 to 11 in the last 12 months is 4.7%, with higher rates among those from impoverished households (Gómez-Restrepo et al., 2016). This gap especially affects vulnerable populations exposed to poverty, mistreatment, early substance use, or contexts of exclusion. In low- and middle-income countries like Colombia, the number of specialized professionals and general staff trained to address mental health is markedly insufficient, which limits the response to the needs of this population (World Health Organization, 2021). This global reality underscores the urgent need to understand and address the societal factors that perpetuate these disparities.

School-based and community-articulated mental health interventions have become key strategies for preventing illness and promoting well-being, aiming to improve mental health literacy, foster help-seeking behaviors, and reduce associated stigma (Bertsia & Poulou, 2023; Campos et al., 2018; Larson et al., 2020; Ma et al., 2023; Matos et al., 2022; Nobre et al., 2021). As part of its Mental Health Action Plan 2013–2030, the WHO proposes adapting and implementing universal school-based interventions focused on promoting mental health, preventing mental disorders, strengthening socioemotional skills, and facilitating early detection and intervention in child and adolescent populations (World Health Organization, 2021).

Resilience refers to the ability to adapt and recover from adversity, maintaining emotional stability and mental health in difficult situations (Bertsia & Poulou, 2023; Bonanno, 2004; Anderson & Priebe, 2021). Compassion, understood as the capacity to recognize and respond to the suffering of others (Peters & Calvo, 2014), has been linked to decreased levels of anxiety and depression, as well as to improvements in general mental health and emotional well-being (Avendaño-Vásquez et al., 2021; Dray et al., 2017; Gilbert, 2019; Gilbert, 2020; Lara-Cabrera et al., 2021; Windle, 2011). These constructs align with the emphasis placed by the WHO Mental Health Action Plan on strengthening protective factors and promoting psychological well-being through preventive and promotive approaches. Together, these skills foster the development of autonomy, a sense of responsibility, an optimistic attitude, and effective social competencies in students (Matos et al., 2022; Mullen et al., 2021; Srikala & Kishore Kumar, 2010). In the case of educators, it has been associated with better emotional regulation, greater empathy, and an enhanced ability to understand others’ affective states (Zhang & Luo, 2023). As a result, resilient teachers can establish more meaningful bonds with their students, especially when they demonstrate emotional sensitivity and understanding regarding student difficulties, while simultaneously promoting the development of other complementary skills (Brooks & Goldstein, 2008).

In low- and middle-income countries, such as Colombia, school-based mental health interventions that promote resilience and compassion have become effective and accessible strategies for fostering emotional and behavioral well-being in adolescents, especially in contexts affected by armed conflict. Teachers, with continuous training and support, can effectively implement these interventions (Fazel et al., 2013; González Ballesteros et al., 2021).

In school contexts marked by high psychosocial vulnerability, the “Conmigo, Contigo y Con Todo” (3C) strategy has become a psychoeducational intervention model aimed at strengthening the resilience, compassion, and mental health of teachers and students. Inspired by cognitive-behavioral therapy and third-generation approaches, the model promotes the development of life skills such as self-knowledge, empathy, emotional regulation, assertive communication, critical thinking, problem-solving, and compassion (González Ballesteros et al., 2021).

While the effectiveness of this type of intervention in strengthening socioemotional well-being has been documented, most studies have focused on expected results or the model’s efficacy, without deeply exploring how these strategies are integrated into the daily lives of children and adolescents in rural and vulnerable areas, what tensions they face, and what learnings emerge during their implementation. As noted in a recent scoping review, reporting on implementation processes is often inconsistent and superficial, and few studies provide detailed accounts of local adaptation or the contextual challenges faced during delivery (Harte & Barry, 2024). This study’s principal objective is to explore children’s and adolescents’ perceptions of the appropriation of the 3C model and how it shapes their understanding of resilience and compassion within their specific educational contexts in rural Boyacá, Amazonas, and Vaupés. Appropriation in public health research refers to the process of adapting and applying concepts or tools in new contexts. It involves the cognitive interpretation of new opportunities for action (Salovaara, 2008) and can be applied to research outcomes (Kensah & Groen, 2008), clinical trials (McDougall et al., 2016), and management tools in healthcare (Côté-Boileau et al., 2020). The process of appropriation involves multiple dimensions, including knowledge acquisition, adaptation, control, and psychological ownership (Mifsud et al., 2015), and can be influenced by sociomaterial and institutional factors (Côté-Boileau et al., 2020). Through a qualitative approach, this study documents how participants experience and transform the model within the school setting, offering insights into its local meaning and practical relevance. As a secondary objective, the study also seeks to analyze participants’ lived experiences with the 3C model to better understand how it becomes integrated into their everyday educational practices.

## Methods

### Study Design

This qualitative study was conducted between June 2023 and December 2024, based on a social heuristic that was constructed through dialogue and broad interpretation of individual realities as narrated by the participants. Through this approach, the perceptions of children and adolescents from 28 educational institutions on resilience, compassion and inclusion were explored, as well as the subjective changes observed at the end of their participation in the psychoeducational model 3C and three months later, in comparison with their initial appraisals. This methodology made possible a rich and detailed description and interpretation of their perceptions, understandings and experiences (Neergaard et al., 2009; Sandelowski, 2000; Vaismoradi et al., 2013).

### Participants and Setting

The study was conducted in collaboration with the Secretariats of Education in Boyacá, Amazonas, and Vaupés, which identified 28 public educational institutions interested in participating in a school-based mental health promotion strategy. For the purpose of this qualitative article, a convenience sample of children and adolescents from the intervention group schools in these three departments was invited to participate in in-person focus groups (FGs). Convenience sampling was used due to logistical constraints and the need to recruit participants directly from schools where the intervention was implemented. Nonetheless, efforts were made to ensure diversity by selecting participants across different ages, genders, and cultural backgrounds within each region.

Inclusion criteria for children and adolescents included being students from 2nd to 11th grade, aged between 6 and 18 years, and being enrolled in the selected schools. Given that children and adolescents as young as 6 years of age participated, their inclusion in the study was initially decided by their parents or legal guardians by signing an informed consent form, who received clear explanations about the objectives, risks and benefits of the project. In addition, each minor participant was asked to give their consent, ensuring that the invitation was made in a language adapted to their age and context, guaranteeing that they understood the purpose of their participation and could freely accept to participate in the focus groups. Exclusion criteria were lack of interest in participating or having a native language other than Spanish. While teachers were trained as multipliers, this article specifically analyzes information from children and adolescents of the intervention group, in line with the study’s objective to explore the appropriation of the 3C model by this population. The study aimed for maximum variation sampling to identify and analyze a wide range of expressions and presentations of the phenomenon, explaining the conditions and contexts of its development.

### Intervention: The “Conmigo, Contigo y Con Todo” (3C) Model

The 3C model is a psychoeducational intervention developed by Fundación Saldarriaga Concha, grounded in resilience and compassion as pathways to inclusion. Designed for various population groups, it is based on cognitive-behavioral principles and structured into 12 modules that address life skills such as self-knowledge, emotional regulation, self-compassion, empathy, interpersonal communication, critical thinking, and decision-making. The methodology includes six key pedagogical moments per session, combining experiential learning and personal reflection. Implementation followed a “train-the-trainer” model: teachers received eight in-person sessions over six months and then replicated the modules with students, supported by follow-up and feedback from facilitators.. The full, detailed description of the strategy is available in a prior publication (Fundación Saldarriaga Concha, 2023).

### Data Collection

Focus groups (FGs) were the primary method for qualitative data collection, conducted at three time points: before the intervention (baseline), six months after (endline), and three months later (maintenance phase). A total of 17 psychologists and 1 speech therapist, all trained in qualitative data collection methods, facilitated these FGs… They used a thematic guide to promote discussion on perceptions and behaviors associated with resilience, compassion, and participants’ views on the 3C strategy. The guide was developed collaboratively by the research team and included a total of 13 questions, organized around the three initial categories of analysis: resilience capacity, compassion capacity, and the 3C strategy. Clear instructions were also provided for conducting the focus groups. The questions were adapted to the age and sociocultural context of the participants to ensure the use of accessible language and references to everyday situations, without compromising the analytical categories. Cultural adaptations were carried out by the group facilitators and were particularly important in Vaupés and Amazonas, given the ethnic diversity of these regions. All focus groups were conducted in Spanish, and transcripts were analyzed in their original language without translation..

For this article, a total of 335 focus groups were conducted with students: 122 in the initial phase, 110 in the final phase, and 103 in the maintenance phase (Graph 1). Most of these focus groups were conducted in person, and only two were conducted virtually, using the Google Meets platform, due to logistical difficulties in accessing the territories.

**Graph 1:**
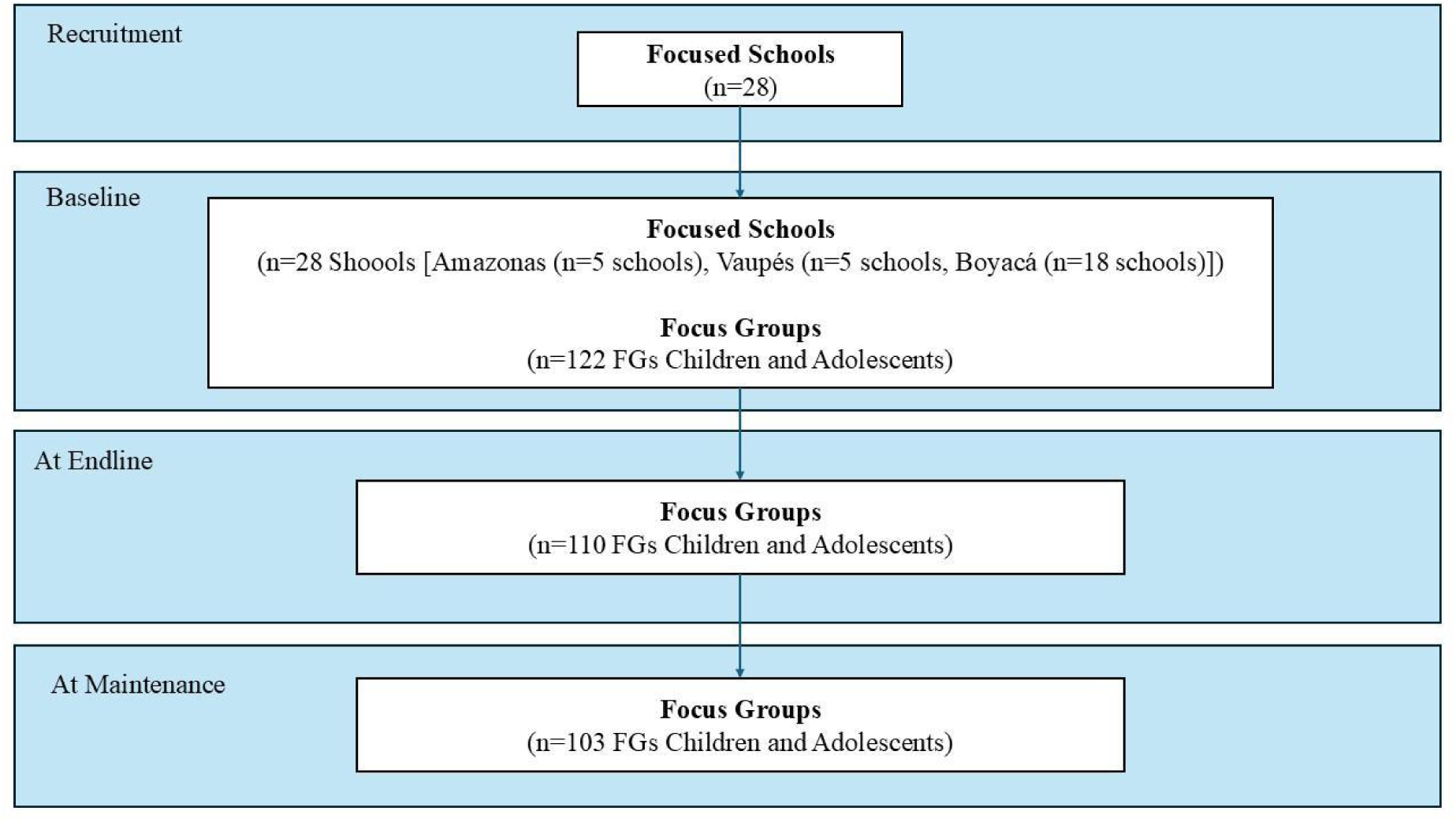
Flow chart of the focus groups conducted before, during, and after the implementation of the “Conmigo, Contigo, Con todo” model. This flow chart illustrates the sequence and timing of the focus groups carried out at three stages of the *Conmigo, Contigo, Con todo* model: Baseline, at Endline, and at Maintenance. The diagram provides a visual representation of how qualitative data collection was structured throughout the process.

### Data Analysis

The content of the focus groups was analyzed using a six-step thematic analysis method, allowing for the identification, organization, and reporting of patterns or themes that guided the understanding and interpretation of participants’ perceptions of resilient and compassionate capacities and observed changes after the intervention. NVivo 14 software (Lumivero, 2023) was used during the deductive coding process, carried out by a sociologist, who employed pre-established categories such as: resilience, compassion, inclusion, and the 3C model. To explore perceptions and changes in resilience and compassion, only these categories were used for this article.

An inductive coding phase was also carried out, which led to the identification of three emergent categories: “Appropriation of the 3C model” (evidence of how children and adolescents internalize and translate the model to their contexts); “Expectations and adaptations” (real adjustments made due to local limitations or needs, comprehension levels, or cultural dynamics); and “Emergent learnings” (personal and professional reflections derived from the implementation process). To ensure coding reliability and enhance the credibility of findings, the initial coding was conducted independently by two researchers. The coding schemes were then compared and refined through iterative discussions until consensus was reached on theme definitions and illustrative segments. Discrepancies were resolved through dialogue, ensuring consistency across transcripts.

Thematic saturation was assessed by territory and study moment (baseline, endline, and maintenance) to ensure information richness and conceptual completeness across diverse contexts. Following the approach outlined by Guest et al. (2020), saturation was operationalized as the point at which the emergence of new codes dropped below 5% across three consecutive focus groups. A cumulative tracking method was applied to monitor the introduction of new codes throughout the coding process.

Across the 335 student focus groups analyzed, saturation was reached after coding 56 groups at baseline, 52 at endline, and 45 at maintenance. Overall, saturation levels exceeded 90% in all phases—93% for baseline, 100% for endline, and 98% for maintenance—indicating that fewer than 7% of codes were newly identified in the final set of transcripts. These results support the adequacy and stability of the thematic structure across all territories.

To enhance interpretative validity, triangulation was applied at three levels: data source (different moments and regions), analyst (multiple coders reaching consensus), and method (combining deductive and inductive coding). Additionally, regular debriefings were held with field researchers to confirm contextual interpretations. This ensured that the findings reflected both thematic breadth and contextual depth.

### Ethical Considerations

The study protocol and all procedures were approved by the Institutional Review Boards (IRB) of Pontificia Universidad Javeriana on May 5, 2022 (Ref. 2022/080). All methods were developed in accordance with relevant guidelines and regulations. The research was classified as minimum risk, as it involved psychological evaluations that did not manipulate individual behaviors and the intervention did not pose risks to mental health. Privacy and confidentiality of individual information were preserved through data de-identification in information systems and databases. A numerical list was used, accessible only to the researcher, linking identification numbers to individuals. Informed consent was obtained from teachers and parents/legal guardians of children and adolescents under 18 years of age, and informed assent was obtained from the minors themselves. Participants were informed of all risks and benefits in clear and simple language, respecting their freedom to participate or withdraw at any time. The authors declare no conflicts of interest. Funding for this project was provided by the Ministry of Science and Technology of Colombia - Minciencias, call 919-2022, Resolution No. 704-2022 (July 14, 2022), Approval Act 39.

Graph 1: Flow chart. Qualitative study.

## Results

The qualitative analysis explored the perceptions and experiences of children and adolescents regarding the appropriation of the “Conmigo, Contigo y Con Todo” (3C) model and its impact on their understanding of resilience and compassion across three rural Colombian departments: Amazonas, Vaupés, and Boyacá. The findings are structured into thematic areas, reflecting the evolution of these concepts from baseline to the maintenance phase.

### 1. Appropriation of the 3C Model: Between Received Knowledge and Lived Experience

Children and adolescents demonstrated a dynamic process of appropriating the 3C model, transitioning from initial exposure to an active integration of its components into their understanding and daily lives. This process was evidenced by how they reinterpreted and applied the model’s principles.

Students’ perceptions of the 3C model components evolved over time. Initially, their understanding was often basic or linked to the general idea of overcoming difficulties. However, in the endline and maintenance phases, students began to explicitly mention the model’s activities and concepts as tools for self-regulation and interpersonal understanding. For example, in Vaupés, students highlighted how “activities and games allowed them to recognize their own capacities and abilities through the gaze of their peers”. This indicates a shift from passive reception to an active translation of the model into their personal and social contexts, demonstrating the internalization of the 3C principles.

### 2. Expectations and Adaptations

At baseline, many students expressed curiosity or a general hope for improvement, often without a clear understanding of what the 3C model entailed. Some even acknowledged unfamiliarity with terms like “resilience”.

As the intervention progressed, students’ expectations shifted from vague hopes to a more concrete understanding of the skills they were acquiring. They described how the model’s activities, particularly the group dynamics, fostered a space for personal reflection and skill development. For example, in Boyacá, students highlighted the 3C model and the “emotions workshop” as a tool that “helped them better understand their emotions”. The flexibility of the model allowed for creative adjustments in its application, as students adapted the learned strategies to their specific challenges and cultural dynamics, which reflects an active reinterpretation of the model in their context.

### 3. Emergent Learnings and Personal Transformations

The intervention fostered significant personal transformations among children and adolescents, leading to new understandings of themselves and their emotional landscapes. This was particularly evident in their self-knowledge, emotional management, and interpersonal relationships, reflecting the application of 3C concepts in their daily lives.

Regarding Self-knowledge, at baseline, students often expressed fear in academic situations and the need to withdraw or seek support when feeling unwell. However, in the maintenance phase, their reflections broadened to include emotional well-being, psychological support, and the ability to analyze situations and relate to others. Students in Amazonas, for instance, learned “the importance of recognizing others’ difficulties and generating empathy”. In Vaupés, students expressed growth in “self-motivation, controlling emotions like anger, setting boundaries, and developing greater responsibility”. These changes demonstrate the internalization of 3C skills.

For Emotional Management, initial strategies were often basic, such as distancing themselves or seeking trusted individuals. Over time, students developed more conscious regulation strategies. In Boyacá, students mentioned new coping forms like “drawing closer to God, playing video games, and taking distance”. The program facilitated a shift from reactive emotional responses to more deliberate and self-aware management, evidencing the application of the model’s tools.

In Interpersonal Relationships, students consistently emphasized the importance of family and social support. In the maintenance phase, their expressions showed a greater emphasis on personal reflection and seeking external support to manage difficult emotions, indicating a deeper integration of relational skills into their coping mechanisms.

### 4. Re-signification of Compassion and Resilience as Part of Model Appropriation

The 3C model facilitated a profound re-signification of resilience and compassion for children and adolescents, moving beyond initial, often limited, understandings to more comprehensive and actionable concepts. This re-signification is a clear indicator of the model’s appropriation.

#### Resilience

##### Amazonas

Students’ initial associations of resilience with resistance or bravery evolved to understand it as “a strength to face adversities, maintain good relationships, and move forward”, demonstrating a reinterpretation of the concept.

##### Vaupés

From associating resilience with confronting problems, students’ understanding broadened to include coexistence, tolerance, and adapting to challenges. In the maintenance phase, some even viewed it as empathy or environmental care, highlighting “putting oneself in others’ shoes”, evidencing a contextualized application.

##### Boyacá

Students progressed from unfamiliarity or linking resilience to resisting pain, to describing it as the capacity to overcome difficult moments, seek solutions, ask for support, and bravely face problems. In the maintenance phase, new aspects were highlighted, such as adapting to life changes, learning from difficult situations, and seeking positive outcomes to overcome challenges, indicating active internalization.

#### Compassion

##### Amazonas

Students’ initial associations of compassion with pity or sorrow evolved to include tolerance, generosity, and emotional help. In the maintenance phase, learnings from the 3C model were mentioned, such as “understanding others’ suffering and not holding grudges, and the importance of respect and support”, showing an application of the model’s principles.

##### Vaupés

Compassion, initially described as giving affection and understanding, evolved towards a more collective sense, highlighting values like empathy, companionship, respect, and kindness, and emphasizing the importance of sharing and living together well.

##### Boyacá

Students’ initial associations of compassion with helping or feeling pity transformed. In the maintenance phase, they reinforced the idea of helping and being supportive, incorporating “the importance of self-love and love for others, valuing and understanding others’ feelings, and differentiating compassion from pity”, reflecting a personal reinterpretation.

#### Empathy

##### Amazonas

Students’ understanding of empathy progressed from supporting and dialoguing to a deeper concept of “putting oneself in others’ shoes,” understanding without judgment, and being a good companion, indicating that the intervention provided tools to help others and that they applied them.

##### Vaupés

Initially, empathy was understood as helping or motivating others. Later, students highlighted the importance of putting themselves in others’ shoes, providing company, comforting, and feeling what others feel, including the idea of being in harmony.

##### Boyacá

Initially, they related empathy to calming emotions, giving advice, supporting, and putting themselves in others’ shoes, highlighting the importance of values and connection with others, in reference to the 3C model. Later, they emphasized empathy as active accompaniment, mentioning the importance of helping find solutions, comforting, advising, and not leaving others alone, which showed an evolution toward a more practical and other-oriented empathy.

#### Self-compassion

##### Amazonas

Initial reflections on unhealed losses and the need for family support evolved into a more well-being-oriented view, with ideas of moving forward, having alone time, crying, and creating positive thoughts to face difficulties.

##### Vaupés

Students’ expressions explicitly linked to the importance of being compassionate with oneself as a strategy to find solutions to problems and better understand others.

##### Boyacá

From mentioning basic coping strategies, students deepened their understanding of identifying emotions and the need to be heard, along with practices like self-forgiveness, patience, meditation, and setting boundaries. In the maintenance phase, a more reflective and learning-oriented view emerged, highlighting “getting up after difficulties, moving forward, distancing themselves from conflicts, and acknowledging mistakes”, demonstrating the application of self-compassion.

## Discussion

This qualitative study provides an in-depth understanding of how children and adolescents in rural areas of Boyacá, Amazonas, and Vaupés appropriated the “Conmigo, Contigo y Con Todo” (3C) model. The findings highlight a dynamic process where participants internalized, reinterpreted, and actively applied the program’s concepts and tools to their unique realities and contexts. As defined, appropriation involves the cognitive interpretation and active integration of new knowledge, skills, or practices into existing frameworks of understanding and daily behaviors, often adapting them to unique contextual realities (Kensah & Groen, 2008; Salovaara, 2008). Initial perceptions of resilience and compassion evolved from abstract or limited understandings to more concrete, actionable, and personally relevant constructs. This nuanced qualitative exploration, focusing on lived experience and the process of appropriation, offers insights that complement traditional efficacy studies by revealing *how* change occurs at the individual and community level, aligning with calls for more theory-driven, data-informed analyses in public health research (Haardörfer, 2019).

The significant evolution in children’s and adolescents’ understanding of resilience demonstrates their active appropriation of the 3C model’s teachings. Initially, resilience was often narrowly perceived as mere resistance or survival in the face of adversity. However, post-intervention, students consistently re-signified resilience as a proactive, multifaceted capacity involving self-knowledge, emotional regulation, and strengthened interpersonal relationships. For instance, in Amazonas, resilience transformed from abstract ideas to “a strength to face adversities, maintain good relationships, and move forward”, indicating an internalization of skills for active coping. Similarly, in Boyacá, the emphasis on adapting to changes and learning from difficult situations highlights the integration of dynamic resilience frameworks, moving beyond individualistic notions to a more relational and adaptive understanding. This appropriation process aligns with ecological and cultural perspectives on resilience, which emphasize its dynamic and socially embedded nature (Bonanno, 2004; Bertsia & Poulou, 2023; L. Theron, 2016). Research on academic resilience further supports the importance of fostering these capacities in school settings (Radhamani & Kalaivani, 2021). Teacher support also emerges as a crucial compensatory factor in rural and disadvantaged contexts, a role fulfilled by the educators in the 3C model (Davidson & Adams, 2013). Children identify factors such as intelligence, family assistance, and school support as promoting academic resilience, reinforcing the multidimensional nature of resilience addressed by the program.

The re-signification of compassion, empathy, and self-compassion observed among children and adolescents further illustrates their successful appropriation of the 3C model. Participants moved beyond a superficial understanding of compassion (e.g., pity or sorrow) to perceiving it as an active, intentional engagement involving support and problem-solving for others. For example, in Boyacá, students differentiated compassion from pity, emphasizing “the importance of self-love and love for others, valuing and understanding others’ feelings”. The shift in empathy towards “active accompaniment” and “helping find solutions” demonstrates the practical application of relational skills learned in the program. This active appropriation of compassion as a skill, rather than just a sentiment, aligns with third-generation cognitive-behavioral approaches that emphasize behavioral commitments and value-driven action, which are foundational to the 3C model (Fundación Saldarriaga Concha, 2023). Compassion can be cultivated in educational settings (Henderson, 2017), and self-compassion during childhood contributes to resilience and empathy in adulthood (Papadimitriou & Karakasidou, 2024), potentially facilitating easier navigation of adolescence (Bluth et al., 2018).

These findings carry significant implications for theory, practice, and policy in public health education. Theoretically, the study underscores the importance of an “appropriation” lens to understand how psychosocial interventions are truly integrated by beneficiaries, especially in culturally diverse and vulnerable contexts. It highlights that resilience and compassion are not just traits to be measured, but dynamic capacities that are re-negotiated and enacted through lived experience. Practically, the results emphasize the value of experiential and culturally adapted approaches, like those in the 3C model, that empower children and adolescents as active agents in their own well-being. The positive shifts in emotional regulation, self-knowledge, and prosocial behaviors suggest that such programs can equip young individuals with crucial life skills for navigating adversity. School-based interventions have shown promise in enhancing socioemotional skills and resilience in rural communities, effectively improving resilience, academic functioning, and emotional regulation (Rich et al., 2022; Westhues et al., 2009). These programs are particularly beneficial for students from economically marginalized communities, addressing barriers to mental health care access (Rich et al., 2022; Stavrou & Kourkoutas, 2017). For policy, these findings provide evidence supporting the continued investment in school-based mental health initiatives within national frameworks, such as Colombia’s National Mental Health Policy (Ministerio de Salud y Protección Social, 2018). The observed appropriation of the model by children and adolescents advocates for integrated, long-term strategies that acknowledge local realities and foster community-led mental health promotion, rather than top-down content delivery. Meta-analyses have confirmed the overall efficacy of school-based interventions in promoting resilience (Cai et al., 2025).

### Strengths and Limitations

Among the strengths of this study is its in-depth qualitative approach, which allowed for a rich exploration of the appropriation process from the perspectives of children and adolescents in an under-researched rural context (Deyra et al., 2020). The use of multiple data collection points (baseline, endline and maintenance) provided a longitudinal view of the evolution of their perceptions, facilitating a broader and contextualized understanding of the phenomenon. Nevertheless, some limitations should be recognized. As a qualitative study, its findings are not generalizable to all populations or contexts. The dynamics of the focus groups, while providing rich information, may have been affected by social desirability bias or peer influence. Also, while the process of ownership of the 3C model is documented, its scope or depth is not quantified, which would require quantitative or complementary mixed methods approaches. In addition, resilience, being a multidimensional construct, requires consideration of cultural components and contextual variations that may have influenced the experiences collected. Gender differences may also affect resilience patterns, but this factor was not specifically analyzed in the present study. These findings underscore the importance of understanding not only the expected outcomes of school-based mental health interventions, but also how children and adolescents interpret, adapt, and actively incorporate these strategies into their daily lives, especially in rural or culturally diverse contexts (Harte and Barry, 2024).

## Conclusion

The findings of this qualitative study suggest that, even in contexts of high adversity such as rural Boyacá, Amazonas, and Vaupés, it is possible to cultivate emotional regulation, compassion, and resilience through culturally sensitive and responsive psychosocial interventions. The appropriation of the “Conmigo, Contigo y Con Todo” (3C) model by children and adolescents was a dynamic process through which they internalized, reinterpreted, and actively applied the program’s core concepts, moving beyond initial understandings toward more actionable and personally meaningful constructs. A shift is observed from an individual understanding of these concepts, initially associated with basic emotions, to a more relational and complex perspective that incorporates others, the community, and the broader context. Furthermore, there is a progressive adoption of conscious, intentional, and sustained strategies to navigate challenges and foster both individual and collective well-being. The observed shifts in their understanding and self-application of resilience and compassion highlight the profound influence of contextually adapted, school-based mental health initiatives. This study underscores the critical role that such programs play in fostering resilience and emotional well-being in vulnerable populations by empowering children and adolescents as active agents in their own psychosocial development, thereby contributing to more humanizing and effective educational models in challenging environments. These findings offer a valuable perspective and a baseline for the implementation of mental health promotion strategies aligned with Colombia’s public policy on socioemotional education in schools. However, future research could explore in greater depth the differences in appropriation or changes in the perspectives of resilience and compassion according to gender, ethnic group or educational level, as well as analyze the sustainability of these learnings over time.

## Data Availability

All data produced in the present study are available upon reasonable request to the authors

## Author Contributions

LMGB conceptualized and designed the study, and is the original designer of the 3C intervention. LMGB, CACR, LEM, JCM and IOJ contributed to the original drafting of the manuscript. LMGB and CACR refined the study’s methodology and provided general project supervision. OEGC and LAL led qualitative data collection. LMGB conducted the critical review of the manuscript. All authors (LMGB, CACR, OEGC, IOJ, LEM, JCM, LAL) participated in the interpretation and discussion of the findings.

## Acknowledgements

We extend our sincere gratitude to all who contributed to this study. We thank the local and municipal governments of Boyacá, Vaupés, and Amazonas for their invaluable support and collaboration. Our deepest appreciation goes to the children and adolescents, their teachers, and the principals of the educational institutions who generously shared their time and experiences, making this research possible. We also acknowledge the dedicated efforts of the entire implementation team for their commitment throughout the project.

The authors acknowledge the use of AI-assisted tools (ChatGPT, OpenAI) for language editing and summarizing early drafts. All content decisions were made by the authors.

## References

Anderson, K., & Priebe, S. (2021). Concepts of Resilience in Adolescent Mental Health Research. The Journal of Adolescent Health : Official Publication of the Society for Adolescent Medicine, 69(5), 689–695. 10.1016/J.JADOHEALTH.2021.03.035

Avendaño-Vásquez, C. J., Reina-Gamba, N. C., Daza-Castillo, L. A., & Quarantini, L. (2021). Nursing interventions in children living under armed conflict situations and quality of life: a scoping review. Journal of Pediatric Nursing, 58, 44–52. 10.1016/j.pedn.2020.11.012

Bertsia, V., & Poulou, M. (2023). Resilience: Theoretical Framework and Implications for School. International Education Studies, 16(2), p1. 10.5539/IES.V16N2P1

Bluth, K., Mullarkey, M.C., & Lathren, C.R. (2018). Self-Compassion: A Potential Path to Adolescent Resilience and Positive Exploration. Journal of Child and Family Studies, 27, 3037–3047. 10.1007/s10826-018-1125-1

Bonanno, G. A. (2004). Loss, trauma, and human resilience: have we underestimated the human capacity to thrive after extremely aversive events? The American Psychologist, 59(1), 20–28. 10.1037/0003-066X.59.1.20

Brooks, R., & Goldstein, S. (2008). The mindset of teachers capable of fostering resilience in students. Canadian Journal of School Psychology, 23(1), 114–126. 10.1177/0829573508316597

Cai, C., Mei, Z., Wang, Z., & Luo, S. (2025). School-based interventions for resilience in children and adolescents: a systematic review and meta-analysis of randomized controlled trials. Frontiers in Psychiatry, 16. doi:10.3389/fpsyt.2025.1594658

Campos, L., Dias, P., Duarte, A., Veiga, E., Dias, C. C., & Palha, F. (2018). Is it possible to “Find space for mental health” in young people? Effectiveness of a school-based mental health literacy promotion program. International Journal of Environmental Research and Public Health, 15(7). 10.3390/ijerph15071426

Côté-Boileau, É., Rouleau, L., Denis, J., & Breton, M. (2020). Appropriating Management Tools in Health Care as Legitimate Sociomaterial Work. 10.5465/ambpp.2020.10322abstract

Davidson, S., & Adams, J. (2013). Adversity and internalizing problems among rural Chinese adolescents: The roles of parents and teachers. International Journal of Behavioral Development, 37(6), 530–541. 10.1177/0165025413503421

Deyra, M., Gay, C., Gerbaud, L., Berland, P., & Pizon, F. (2020). Global Health Determinants Perceived and Expressed by Children and Adolescents Between 6 and 17 Years: A Systematic Review of Qualitative Studies. Frontiers in Pediatrics, 8, 115. 10.3389/fped.2020.00115

Dray, J., Bowman, J., Campbell, E., Freund, M., Hodder, R., Wolfenden, L., Richards, J., Leane, C., Green, S., Lecathelinais, C., Oldmeadow, C., Attia, J., Gillham, K., & Wiggers, J. (2017). Effectiveness of a pragmatic school-based universal intervention targeting student resilience protective factors in reducing mental health problems in adolescents. Journal of Adolescence, 57, 74–89. 10.1016/J.ADOLESCENCE.2017.03.009

Fazel, M., Patel, V., Thomas, S., & Tol, W. (2013). Mental health interventions in schools in low-income and middle-income countries. BMC Public Health, 13, 835. 10.1186/1471-2458-13-835

Gilbert, P. (2019). Explorations into the nature and function of compassion. Current Opinion in Psychology, 28, 108–114. 10.1016/J.COPSYC.2018.12.002

Gómez-Restrepo, C., Aulí, J., Tamayo Martínez, N., Gil, F., Garzón, D., & Casas, G. (2016). Prevalencia y factores asociados a trastornos mentales en la población de niños colombianos. Revista Colombiana de Psiquiatría, 45(S1), 39–49. 10.1016/j.rcp.2016.06.010

González Ballesteros, L. M., Flores, J. M., Ortiz Hoyos, A. M., Londoño Tobón, A., Hein, S., Bolívar Rincon, F., Gómez, O., & Ponguta, L. A. (2021). Evaluating the 3Cs program for caregivers of young children affected by the armed conflict in Colombia. Journal on Education in Emergencies, 7(2), 212–252. 10.33682/14b2-4nmm

Guest, G., Namey, E., & Chen, M. (2020). A simple method to assess and report thematic saturation in qualitative research. PLOS ONE, 15(5), e0232076. 10.1371/JOURNAL.PONE.0232076

Haardörfer, R. (2019). Taking Quantitative Data Analysis Out of the Positivist Era: Calling for Theory-Driven Data-Informed Analysis. Health Education & Behavior, 46(4), 537–540. 10.1177/1090198119853536

Harte, P., & Barry, M. M. (2024). A scoping review of the implementation and cultural adaptation of school-based mental health promotion and prevention interventions in low-and middle-income countries. Cambridge Prisms: Global Mental Health, 11, e55. doi:10.1017/gmh.2024.48

Henderson, Deborah J. (2017) Compassion and education: cultivating compassionate children, schools and communities by A. Peterson. Curriculum Perspectives, 37(2), pp. 211–212.

Hollister-Wagner, G.H., Foshee, V.A., & Jackson, C. (2001). Adolescent Aggression: Models of Resiliency. Journal of Applied Social Psychology, 31, 445–466. DOI:10.1111/J.1559-1816.2001.TB02050.X

Kensah, D., & Groen, A. (2008). Appropriation of value in Biomedical research outcome at Public Research Organisations. The Dutch Institute for Knowledge Intensive Entrepreneurship, University of Twente. 10.3990/2.268487997

Kohn, R., Ali, A., Puac-Polanco, V., Figueroa, C., López-Soto, V., Morgan, K., et al. (2018). Mental health in the Americas: An overview of the treatment gap. Revista Panamericana de Salud Pública, 42, e165. 10.26633/RPSP.2018.165

Lara-Cabrera, M. L., Betancort, M., Muñoz-Rubilar, C. A., Novo, N. R., & De las Cuevas, C. (2021). The mediating role of resilience in the relationship between perceived stress and mental health. International Journal of Environmental Research and Public Health, 18(18). 10.3390/ijerph18189762

Larson, K. E., Nguyen, A. J., Orozco Solis, M. G., Humphreys, A., Bradshaw, C. P., & Lindstrom Johnson, S. (2020). A systematic literature review of school climate in low and middle income countries. International Journal of Educational Research, 102, 101606. 10.1016/j.ijer.2020.101606

Ma, K. K. Y., Anderson, J. K., & Burn, A. M. (2023). Review: School-based interventions to improve mental health literacy and reduce mental health stigma – a systematic review. Child and Adolescent Mental Health, 28(2), 230–240. 10.1111/CAMH.12543

McDougall, R., Martin, D., Gillam, L., Hallowell, N., Brookes, A., & Guillemin, M. (2016). Therapeutic appropriation: a new concept in the ethics of clinical research. Journal of Medical Ethics, 42(12), 805–808. 10.1136/medethics-2016-103612

Matos, M., Albuquerque, I., Galhardo, A., Cunha, M., Lima, M. P., Palmeira, L., Petrocchi, N., McEwan, K., Maratos, F. A., & Gilbert, P. (2022). Nurturing compassion in schools: A randomized controlled trial of the effectiveness of a Compassionate Mind Training program for teachers. PLoS ONE, 17(3). 10.1371/JOURNAL.PONE.0263480

Mullen, C. A., Shields, L. B., & Tienken, C. H. (2021). Developing Teacher Resilience and Resilient School Cultures. AASA Journal of Scholarship & Practica, 18(1), 8–24.

Neergaard, M. A., Olesen, F., Andersen, R. S., & Sondergaard, J. (2009). Qualitative description – the poor cousin of health research? BMC Medical Research Methodology, 9(1), 52. 10.1186/1471-2288-9-52

Nobre, J., Oliveira, A. P., Monteiro, F., Sequeira, C., & Ferré-Grau, C. (2021). Promotion of Mental Health Literacy in Adolescents: A Scoping Review. 10.3390/ijerph18189500

Radhamani, K., & Kalaivani, D. (2021). Academic Resilience among Students: A Review of Literature. International Journal of Research and Review, 8(6), 360–369. 10.52403/ijrr.20210646

Rich, B. A., Starin, N. S., Senior, C. J., Zarger, M. M., Cummings, C. M., Collado, A., & Pande, Y. (2022). Improved resilience and academics following a school-based resilience intervention: A randomized controlled trial. Evidence-Based Practice in Child and Adolescent Mental Health, 8(3), 252–268. 10.1080/23794925.2022.2025630

Papadimitriou, A., & Karakasidou, E. (2024). Self-compassionate children, resilient and empathetic adults. International Journal of Environmental Research and Public Health, 21(5), 612. 10.3390/ijerph21050612

Peters, D., & Calvo, R. (2014). Compassion vs. empathy: Designing for resilience. https://www.researchgate.net/profile/Rafael_Calvo/publication/274475960_Compassion_vs_empathy/links/5699807e08aeeea98594927e.pdf

Sandelowski, M. (2000). Whatever happened to qualitative description? Research in Nursing & Health, 23(4), 334–340. 10.1002/1098-240X(200008)23:4

Salovaara, A. (2008). Inventing new uses for tools: A cognitive foundation for studies on appropriation. Human technology : an interdisciplinary journal on humans in ICT environments, 4, 209–228. 10.17011/HT/URN.200811065856

Srikala, B., & Kishore Kumar, K. V. (2010). Empowering adolescents with life skills education in schools-School mental health program: Does it work. Indian J Psychiatr, 52(4), 344–349. 10.4103/0019-5545.74310

Stavrou, P., & Kourkoutas, E. (2017). School Based Programs for Socio-emotional Development of Children with or without Difficulties: Promoting Resilience. American Journal of Educational Research, 5, 131–137.

Theron, L. (2016). Cultural pathways to resilience: African approaches to fostering well-being in children and adolescents. Journal of Psychology in Africa, 26(3), 241–252. 10.1080/14330237.2016.1176258

Ungar, M., Russell, P., & Connelly, G. (2014). School-Based Interventions to Enhance the Resilience of Students. Journal of Educational and Developmental Psychology, 4, 66. DOI:10.5539/JEDP.V4N1P66

Vaismoradi, M., Turunen, H., & Bondas, T. (2013). Content analysis and thematic analysis: Implications for conducting a qualitative descriptive study. Nursing & Health Sciences, 15(3), 398–405. 10.1111/NHS.12048

Westhues, A., Hanbidge, A.S., Gebotys, R.J., & Hammond, A. (2009). Comparing the Effectiveness of School-Based and Community-Based Delivery of an Emotional Regulation Skills Program for Children. School Social Work Journal, 34, 74–95.

Windle, G. (2011). What is resilience? A review and concept analysis. Reviews in Clinical Gerontology, 21(2), 152–169. 10.1017/S0959259810000420

World Health Organization. (2021). Comprehensive mental health action plan 2013–2030. World Health Organization. https://www.who.int/publications/i/item/9789240031029

World Health Organization. (2021). Mental health atlas 2020. Geneva: World Health Organization. https://www.who.int/publications/i/item/9789240036703

Zhang, S., & Luo, Y. (2023). Review on t§he conceptual framework of teacher resilience. Frontiers in Psychology, 14, 1179984. 10.3389/fpsyg.2023.1179984

